# Effects of Metformin compared to Thiazolidinediones on vascular endothelial function in Polycystic Ovary syndrome: A systematic review

**DOI:** 10.1101/2020.01.23.20018580

**Authors:** Hoimonty Mazumder, Farah Faizah, Md. Mahbub Hossain

## Abstract

Metformin and Thiazolidinediones are insulin sensitizers used as the first-line treatment of PCOS, which are recently being examined for their extensive beneficial impacts at biochemical and molecular level of body function in Polycystic ovary syndrome. Therefore, purpose of this review is to compare the effectiveness of Metformin and Thiazolidinediones on vascular endothelial function in PCOS. MEDLINE (1966- October 2018), CINAHL (1982 to October 2018), Cochrane Central Register of Controlled Trials and Cochrane Menstrual Disorders and Subfertility Group trials register (October 2018) and Google Scholar were searched electronically without language restriction. Three Randomized controlled trials (RCTs) were selected with a total of 90 participants comparing effect of Metformin (Intervention) to Thiazolidinediones (Comparator) with an outcome on endothelial function among women with PCOS. In results, Meta-analysis shows no statistically significant difference on endothelial-dependent vasodilation between Metformin and Thiazolidinedione (p value= 0.59 at 95% CI - 1.31, 0.74).

## 1. Introduction

Polycystic ovary syndrome is a complex disorder of altered reproductive and metabolic function, affects 2-20% of women in the reproductive age group (Baio, 2017). Prevalence of Polycystic ovary syndrome varies across the world because of contentious diagnostic criteria; albeit as per Rotterdam criteria 20% of women suffering from this disorder (Rotterdam & Group, 2004; Society et al., 2010). PCOS is equally prevalent in developed and developing parts of the world precipitated by rapidly changing socioeconomic and lifestyle pattern substantially created a great burden in the entire course of life. In India, 41% women struggled with Polycystic ovary syndrome where young women constitute the highest share (37.1%) (Choudhary, Jain, & Chaudhari, 2017). It was primarily reported as reproductive endocrinopathy, but recent studies indicate an increased risk of developing cardiovascular diseases with further correlation with metabolic alteration. Hence, clinical manifestations of metabolic disorder, including insulin resistance, dyslipidemia, high blood pressure and type-2 diabetes mellitus (C J G Kelly, Connell, Cameron, Gould, & Lyall, 2000) are frequently present in Polycystic ovary syndrome potentially able to exert the risk of cardiovascular diseases (Després & Marette, 1994; Mykkänen, Haffner, Rönnemaa, Bergman, & Laakso, 1997). Cardiovascular disease is the leading cause of mortality and morbidity in women, including the estimated risk of developing CVD amongst women is 39% (Coulter, 2011; Shedd & Limacher, 2017). A hallmark of attributable risk factors of vascular alteration induced by metabolic and hemodynamic divergence eventually led serious cardiovascular complications in Polycystic ovary syndrome (Ginsberg, 2000; Christopher J G Kelly et al., 2002).

Endothelial dysfunction refers to precocious structural and functional arterial changes significantly attributable in atherosclerotic progress (Christopher J G Kelly et al., 2002; Paradisi et al., 2001; E. O. Talbott et al., 2000). It is an early feature of vascular pathology (Bonetti, Lerman, & Lerman, 2003; E. Talbott et al., 1995) collectively manifests by impaired arterial vasodilation, increased arterial stiffness, and increased vascular oxidative stress (Bonetti et al., 2003). Though there are numerous controversies regarding endothelial function in PCOS, however, some recent studies significantly demonstrated impaired endothelial function among women with PCOS (Christopher J G Kelly et al., 2002; Kravariti et al., 2005). Endothelium is a single layer of cell lining the internal surface of blood vessels plays an essential role in the regulation of vascular homeostasis and also acts as an endocrine organ (Bonetti et al., 2003; Cardiologist, 2002). The features of metabolic aberration in PCOS includes insulin resistance, dyslipidemia, hypertension, and diabetes causes increased oxidative stress which substantially involved in the pathogenesis of endothelial dysfunction and promotes cellular damage (Cai & Harrison, 2000; Tomasian, Keaney Jr, & Vita, 2000). Diminished bioavailability of Nitric Oxide in counter response promotes plasma concentration of inflammatory mediators, thus contributing to atherogenesis (Bonetti et al., 2003; Ilie et al., 2010).

Metformin is a biguanide which is pharmacologically indicated to improve insulin sensitivity in the treatment of type-2 Diabetes Mellitus. It is widely prescribed for patients with PCOS due to its insulin-mediated beneficial effects in Polycystic ovary syndrome. Several clinical trials suggested significant improvement in vasodilatation (Diamanti-Kandarakis et al., 2005; Jensterle et al., 2008; Orio et al., 2005; Romualdi et al., 2008) whereas few others have concluded negatively (Lowenstein, Damti, Pillar, Shott, & Blumenfeld, 2007; Luque-Ramírez, Mendieta-Azcona, del Rey Sánchez, Matíes, & Escobar-Morreale, 2009; Meyer, McGrath, & Teede, 2007). Likewise, Thiazolidinediones (Rosiglitazone and Pioglitazone) has shown beneficial effects on the microvasculature, including improved endothelial function, decreased inflammatory markers, and reduce blood pressure along with its typical function on glycemic control and decrease insulin resistance (M. Chen & Lee, 2004; Nakamura et al., 2004).

Since, patients with PCOS are more likely to develop CVD because of increased biomarkers and both insulin sensitizers (Metformin and Thiazolidinediones) are widely used in the treatment of PCOS empirically, it is essential to examine the roles of these commonly used drugs for PCOS in improving endothelial function. To our knowledge, there is no systematic review on the use of metformin and thiazolidinediones to see their effect on endothelial function in PCOS-affected population. This review contributes to this knowledge gap. The objective of this review is to systematically evaluate the current evidence on comparative effect of metformin and thiazolidinediones on endothelial function from Randomized controlled trials (RCTs).

## 2. Methods

The population was women with PCOS where diagnosis was done based on hyperandrogenism, menstrual dysfunction (Oligo-ovulation or anovulation), hirsutism, acne and/or ultrasound findings of Polycystic ovaries. Here, metformin was compared to thiazolidinediones (Pioglitazone and Rosiglitazone) to evaluate their effect on endothelial vasodilation. This review focused on a very specific outcome-endothelial function which is considered as an important predictor of cardiovascular pathology.

All randomized control trials (RCTs) were reviewed comparing metformin with Thiazolidinediones. In regards, two thiazolidinediones (Pioglitazone and Rosiglitazone) were considered, the other one (Troglitazone) proscribed because of its fatal effect on human body. Non-RCTs, Crossover trials and quasi-randomized trials were excluded. The literature search had no time or language constraint and searching was run till 30/10/2018. MEDLINE (1966-October 2018), CINAHL (1982 to October 2018), Cochrane Central Register of Controlled Trials and Cochrane Menstrual Disorders and Subfertility Group trials register (October 2018) and Google Scholar were searched electronically. Reference list of included studies and other relevant review articles were checked for any additional citation. MeSH terms and other free term were used as keyword to retrieve more relevant articles. Search terms were “Polycystic ovary syndrome” or “PCOS” and “metformin” or “biguanides” and “thiazolidinediones” or “Rosiglitazone” or “Pioglitazone” and “endothelium” or “endothelial function”.

### 2.1. Study selection and quality assessment

All retrieved studies were screened for relevance of review objectives and quality in methodology. Study selection was conducted by two reviewers by applying inclusion and exclusion criteria. Data extraction also undertaken by the same reviewers independently and assessed. Any disagreements were resolved consensually. Only, randomized control trials have chosen for this review because of its top hierarchical position in terms of quality research. It is coined as ‘gold standard of scientific treatment evidence’ that led to considering in inclusion criteria of this study. Hence, Randomization considered as the only approach to prevent systematic differences in baseline characteristics among two or more intervention groups (Green & Higgins, 2005). Moreover, all included studies have a valid sample and comparing groups being randomized from the same population. Intervention includes single administration of Metformin without any adjunct therapy and Thiazolidinediones (Pioglitazone and/or Rosiglitazone) as comparator. Women of reproductive age group diagnosed with Polycystic ovary syndrome are the population in this review.

**Figure 1.**
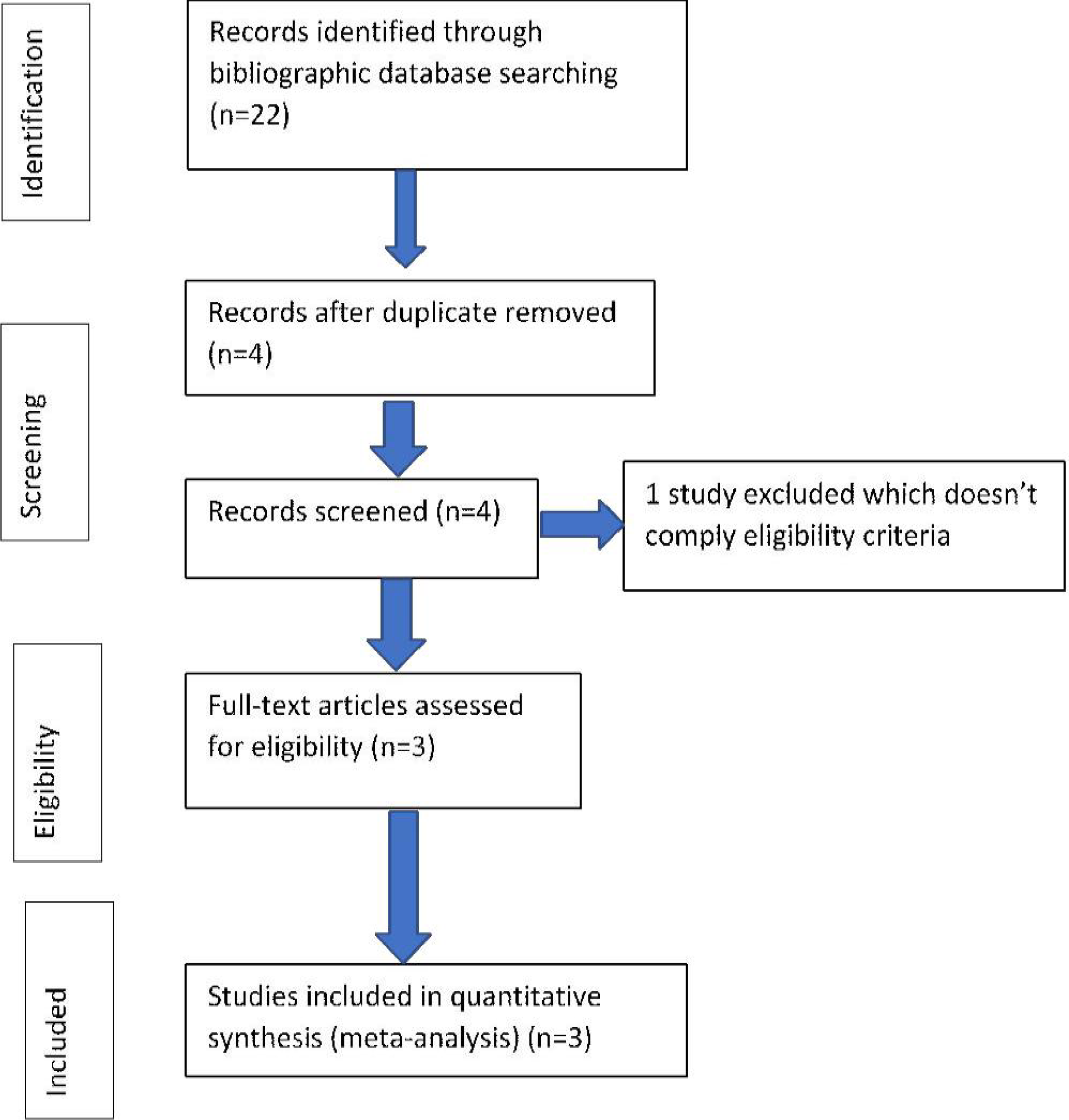
Flow diagram showing the number of publications identified in the literature search.

The diagnosis of PCOS in all three trials required at least two of the following three features: (i) menstrual dysfunction (oligomenorrhoea or amenorrhoea), (ii) clinical or biochemical hyperandrogenism and (iii) polycystic ovaries on ultrasound (Morin-Papunen et al., 2000, 2003; Harborne et al., 2003b; Rautio et al., 2005). The duration of trials ranged 3 to 6 months. Two out of three included trials, dosage of metformin was 850 mg twice daily for 6 months (Naka et al., 2010; Jensterle et al. 2008) whereas 500 mg metformin was applied twice daily for 3 months in the rest one (Mohiyiddeen et al., 2013). As a comparator, Rosiglitazone was (4 mg once daily) used in two studies; amongst, one for 6 months and the other one lasts for 3 months (Jensterle et al. 2008; Mohiyiddeen et al., 2013). Only one trial assigned pioglitazone 30 mg single dose for 6 months (Naka et al., 2010). The total 98 people was enrolled in to the selected trials amongst 8 patients were either lost from the follow-up or excluded before randomization. Therefore, a total of 90 patients with mean age range for individual trials 23.1+/- 3.7 to 30+/-0.9 were randomized by computer-generated system for interventions. All three trials were enrolled fewer than 50 participants in hospital either from department of Gynaecology or Endocrinology. Assessment of endothelial function as review outcome was measured by flow-mediated dilatation in two RCTs (Naka et al., 2010; Jensterle et al. 2008) whereas one trial used acetylcholine induced endothelial-dependent vasodilatation (Mohiyiddeen et al., 2013) performed by doppler imager. Though, all included studies were randomized controlled trial however, two of these designed as open label (Naka et al., 2010; Mohiyiddeen et al., 2013) while one trial didn’t specify about (Jensterle et al. 2008).

Quality assessment of included studies was conducted by employing the Critical Appraisal Skills Programme (CASP) and Cochrane recommended ‘Risk of Bias toolkit hence, renders specific guidelines regarding the high, low and unclear risk of bias thereby selection bias, performance bias, detection bias, attrition bias, reporting bias and other biases could have been evaluated.

### 2.2. Data Extraction

As per PRISMA checklist, data extraction was carried out and presented. A pre-designed codebook was used to extract data on study characteristics, intervention design, and outcomes assessment.

### 2.3. Statistical analysis

Endothelial function in all selected trials was measured by two different tools and represented as baseline and follow-up data. For continuous data, the mean baseline and post-intervention data were measured by RevMan software (Version 5.3) and weighted mean differences with 95% CI were calculated. When the same continuous data were measured by different scales then mean +/- standard deviation were measured for each group and standardized mean differences were calculated with 95% CI. Heterogeneity (I2) was determined by inspecting the results of different studies how sporadically distributed. Statistically, chi-squared test *P* 0.05 represents homogeneity. An *I*^*2*^ value greater than 50% indicated heterogeneity between studies.

## 3. Results

### 3.1. Study characteristics

A total sixteen potentially relevant studies were identified initially through title searching. Of these, four studies were included following de-duplication. De-duplication was run in Endnote software to avoid double counting and overestimation of the effect (Wager & Wiffen, 2011). Finally, three studies were chosen subsequently following abstract and full-text review by applying review eligibility criteria. Eighty studies were excluded because of lack of randomization, failure to meet inclusion criteria, deploying drug combination as intervention or studies conducted in a different group of population other than PCOS. The homogenous characteristics of included studies in terms of population, intervention, comparator and outcome led those studies to be included. The detailed characteristics of the finally recruited studies are presented in Table 1.

**Table 1:** Characteristics of the recruited studies.

| Author and year of Publication | Country of study | Population | Sample size | Study type | Age of Participants |  | Study setting and recruitment criteria | Dose and frequency of intervention drug |  | Clinical method of measuring vascular diameter | Loss to follow up | Outcome |
| --- | --- | --- | --- | --- | --- | --- | --- | --- | --- | --- | --- | --- |
|  |  |  |  |  | Intervention group | Comparison Group |  | Intervention group | Comparison group |  |  |  |
| Jensterle et al. 2008 | Slovenia | Women With Polycystic ovary Syndrome | 28 | RCT; follow up at 6 months | Mean age in Intervention group 23.1 ± 3.7 years | Mean age in comparison group 25.0 ± 4.9 years | Department of Endocrinology, University Medical Centre, Slovenia; National Institute of Child Health and Human Development criteria | Metformin 850 mg twice daily | Rosiglitazone 4 mg once daily | Flow-mediated dilation (FMD) of the Brachial artery | Two women Were excluded due to - protocol Violation | Endothelial dependent dilatation of brachial artery |
| Naka et al., 2010 | Greece | Women With Polycystic ovary syndrome | 30 | RCT; follow up at 6 months | Mean age 23.3 ± 4.9 years | Mean age 23.3 ± 4.9 years | Department of Endocrinology at the University Hospital of Ioannina, Greece; Patient recruited with PCOS who has two of following three features oligo-ovulation or anovulation, Hyperandrogenism, and Polycystic Ovaries. | Metformin 850 mg twice daily | Pioglitazone 30 mg once per day | Flow-mediated dilation (FMD) of the brachial artery | One woman in the Pioglitazone group was Lost | Endothelial-dependent dilatation of brachial artery |
| Mohiyiddeen et al., 2013 | United Kingdom | Women with Polycystic ovary Syndrome | 50 | RCT; follow up at 3 months | Mean age of Intervention group $30.0 \pm 0.9$ years | Comparison group $29.0 \pm 1.0$ years | Department of Gynecology, Tameside General Hospital, Ashton-Under-Lyne, UK; Rotterdam consensus of PCOS including hyperandrogenism, chronic anovulation, and polycystic ovaries | Metformin 500 mg twice daily | Rosiglitazone 4 mg once daily | Perfusion-Measurement for Acetylcholine obtained by laser Doppler imager | Ten patients Were excluded because of Their intention to conceive. Three patients in The Metformin group and 2 patients in the Rosiglitazone group | Endothelial-dependent dilatation for the delivery of Acetylcholine |

### 3.2. Quality assessment of the included studies

Since All included RCTs followed open-label design, so blinding of participants and allocation concealment didn’t perform subjected towards the high risk of bias. Kahan et al., 2014 hence, argued that in case of open label randomized trials, adopting a set of criteria or definition to ensure study objectivity prior recruitment thus able to reduce the potential impact of unblinded assessment. However, assessors of all three studies who carried out baseline and follow-up clinical investigations were blinded regarding randomization of interventions that minimized likelihood of result to be biased. It is suggested for all open label RCTs that to keep blinding outcome assessment ensured bias reduction effectively.

All the included studies in this review were assigned to interventions following randomization, done by using a computer-generated system. Despite having such great flaws, these open label studies were considered in this review because of some reason stated follows-

1. After reviewing these studies, it was clear that Researcher accepted a definition or set of criteria for diagnosing the Polycystic ovary syndrome prior recruitment and randomization. This attempt can reduce the potential impact of unblind assessment (Kahan et al., 2014).
2. “Endothelial function” is considered as primary outcome, therefore unlikely to introduce bias (Kahan et al., 2014). Moreover, the Endothelial function required several technical assistances to be evaluated therefore concealment and blinding seemed unfeasible.
3. Blinding of outcome assessment ensured further minimizing the risk of bias of included studies. Assessors in all 3 studies who involved in investigating outcomes were blind which reduces the likelihood of results to be biased.

Finally, attrition bias refers to the incomplete reporting of outcome data (Higgins et al., 2011) which was absent within selected studies. Selective reporting and other biases also considered in the appraisal of studies as per the guidance of Cochrane ‘Risk of Bias’ tool.

### 3.3. Intervention, study design, and outcomes

All studies were conducted in a hospital setting, participants underwent in the process of randomization and finally intervention and comparator were assigned for a defined time period. The duration of interventions of included trials ranging from 3 to 6 months and dosage of Metformin ranging from 500 to 850 mg twice daily. As a comparator this review considered Thiazolidinediones while two trials had used Rosiglitazone 4 mg for 3 and 6 months and a single trial assigned Pioglitazone 30 mg once daily for 6 months. The included studies randomized a total of 90 women, two of these randomly assigned interventions (Metformin) and Comparator (Pioglitazone or Rosiglitazone) while other one randomized into three groups (Metformin, Pioglitazone and placebo/no treatment group). The number of participants in each group, their baseline and end of the intervention evaluations are presented in Table 2.

**Table 2:** Intervention and outcome evaluation.

| Study | Study Population Completed follow-up | Intervention (Metformin) |  | The comparator (Thiazolidinedione) |  | P value |  |
| --- | --- | --- | --- | --- | --- | --- | --- |
|  |  | Baseline | After intervention | Baseline | After intervention | Statistical significance for Metformin (expressed in p value) | Statistical significance for Thiazolidinedione (expressed in p value) |
| Jensterle et al. 2008 | Intervention, n= 15<br>Comparator, n=11 | 4.2 ± 6.6 | 10.2 ± 5.9 | 2.9 ± 3.2 | 7.6 ± 4.9 | 0.036 | 0.026 |
| Naka et al. 2010 | Intervention, n= 15<br>Comparator, n= 14 | 4.07 ± 2.66 | 9.33 ± 2.20 | 4.43 ± 2.61 | 9.32 ± 3.14 | <0.001 | <0.001 |
| Mohiyiddeen et al. 2013 | Intervention, n= 17<br>Intervention, n=18 | 0.58 ± 0.03 | 0.63 ± 0.04 | 0.62± 0.05 | 0.69± 0.05 | 0.34 | 0.5 |

Jensterle et al. (2008) randomized 28 women diagnosed Polycystic ovary syndrome and administered Metformin (n=15) and Rosiglitazone (n= 13) in to two groups. Likewise, Naka et al., 2011 recruited 43 PCOS patients in a University Hospital out-patient department was randomized into three different arms: no treatment (n=14), Metformin 850 mg (n=15) and Pioglitazone 30 mg (n=14) for 6 months. Since this review aims to find out the comparative efficacy of Metformin with Thiazolidinediones, therefore, two arms (Metformin and Pioglitazone) were considered for meta-analysis. The last RCT (Mohiyiddeen et al., 2013) enrolled a total of 50 patients with PCOS in hospital setting based on criteria of Rotterdam consensus. Following recruitment, 10 patients were excluded because of their intention to conceive. Remaining 40 patients were randomized into two groups by using the computer-generated system. Total 35 patients completed the follow-up and analysis where 17 women received Metformin 500 mg twice daily and 18 women received Rosiglitazone 4 mg once daily for 3 months. In order to assess the significance of endothelial function baseline and endpoint data were recorded by Flow-mediated dilation (FMD) of the brachial artery scanned by using a high-resolution advance ultrasound system (Jensterle et al. 2008; Naka et al., 2011) whereas Mohiyiddeen et al. (2013) applied non-invasive measurement of skin perfusion following induction of acetylcholine in which internal circumference of arterial chamber was recorded by Doppler imager.

### 3.4. Meta-analysis

The primary outcome was endothelial-dependent vasodilatation measured by Flow-mediated dilatation in two studies (Jensterle at al., 2008; Naka et al., 2011) and only one used acetylcholine induced assessment of endothelial-dependent dilatation (EDD) (Mohiyiddeen et al.,2013). Meta-analysis demonstrated no significant difference in endothelial-dependent vasodilation between Metformin and Thiazolidinediones (SMD -0.28, P=0.59, 95% CI -1.31 to 0.74) as shown in Figure 2. The standard mean difference (SMD) is used when pooling continuous data on outcomes measures by different scales. Heterogeneity was tested using chi-squared test and I_2_, Chi_2_= 11.15, I_2_= 82%.

**Figure 2:**
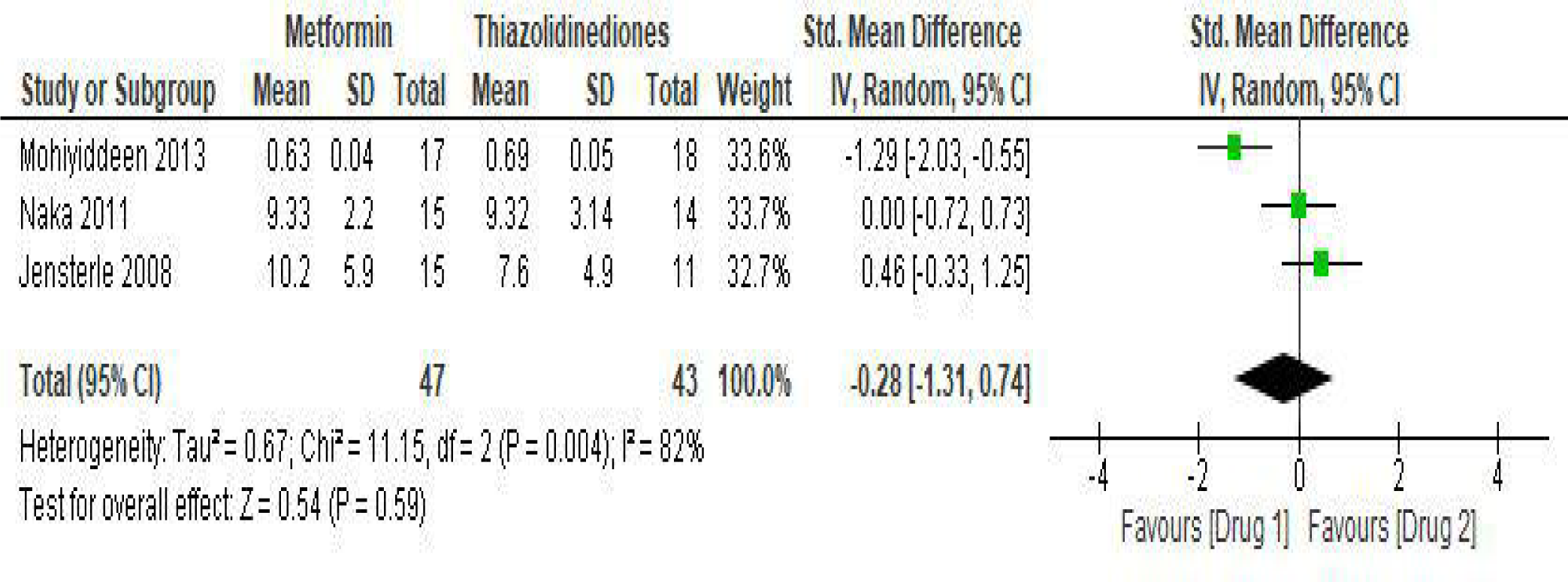
Comparison of Metformin versus Thiazolidinediones with outcome of Endothelial function.

## 4. Discussion

Since, Polycystic ovary syndrome and its other comorbidities eventually predispose cardiovascular pathology (Lowenstein, Damti, Pillar, Shott, & Blumenfeld, 2007) therefore, it is important to take this issue under consideration during clinical management of PCOS. Hence, Metformin is one of the preferred choices of clinicians in treating insulin resistance and Thiazolidinediones also used to some extent for the same purpose. Improvement in cardiometabolic syndrome largely observed following metformin therapy by lowering systolic blood pressure and LDL indicates its cardio-protective nature (Lord et). On the other hand, Thiazolidinediones improve endothelial dependent vasodilation and in counter-response attenuated vasoconstriction through cell-mediated vascular regulation (Kotchen, Zhang, Reddy, & Hoffmann, 1966; Zhang, Sowers, Ram, Standley, & Peuler, 1994). In addition, Pioglitazone significantly improves insulin sensitivity and reducing abnormal lipid profile or inflammatory markers, thus preventing progress of atheromatous change on microvasculature (Berhanu et al., 2006; Szapary et al., 2006).

This review has found that up to 6 months of treatment with Metformin compared with Thiazolidinediones in PCOS is associated with no significant difference in vascular function. There are very few randomized control trials explored the effects of Metformin and Thiazolidinediones on endothelial function in PCOS, but numerous controversies exist regarding the effect of Metformin on vascular endothelium. Besides, Thiazolidinediones showed lack of presence in experimental studies to identify its effects on endothelium. Two out of three included studies showed significant improvement of endothelial function following 6 months administration of Metformin and Thiazolidinediones (Jensterle et al., 2008; Naka et al., 2011) amongst one study found Metformin as effective as Rosiglitazone (p =0.045) (Jensterle et al., 2008) and other one observed both effects comparable (Naka et al., 2011). Similar findings were observed in a clinical trial with significant improvement of Flow-mediated dilation (FMD) following 6 months of Metformin therapy in 30 young, normal weight women with PCOS (Orio et al., 2005). Besides, Rosiglitazone, though data is more limited however, Tarkun et al. (2005) shown to improve vascular function and Pioglitazone, as well, exerts significant increase in endothelium-dependent vascular response (Luque-Ramírez, Mendieta-Azcona, Álvarez-Blasco, & Escobar-Morreale, 2009; Papathanassiou et al., 2009). In contrast, Mohiyiddeen et al. (2013) deduced no significant change on endothelial-dependent vasodilatation following 3 months administration of Metformin and Rosiglitazone but endothelial-independent function had improved in the Rosiglitazone group. Short duration of intervention (3 months) and low dosage of Metformin therapy might be the reason of insignificant outcome of this RCT (Mohiyiddeen et al., 2013). Several clinical trials with 850 mg of Metformin bid for 6 months demonstrated improved endothelial function amongst Polycystic ovary syndrome (Diamanti-Kandarakis et al., 2005; Orio et al., 2005). Hence, longer periods of Metformin administration may show a significant trend of improvement in vascular response. Also, Tarkun et al. (2005) reported beneficial effects on endothelial function in PCOS following 12 months treatment duration. Similarly, Naka et al. (2011) shown positive vascular response after 6 months of trial with Pioglitazone (Naka et al., 2011). Future biomedical and clinical research in this domain should explore how administering Metformin and Thiazolidinediones can potentially impact patho-physiological systems beyond their primary indications for PCOS. Moreover, prospective research should evaluate how dose-response relationships between administration of these drugs and endothelial function can be examined through stronger study designs with adequate statistical power and larger sample size.

## 5. Limitations of the review

Although, all the included RCTs were randomized but the design of the study was open label which has the possibilities to introduce potential bias. According to Cook, et al., (1993) inclusion of unpublished trials is always controversial, in contrast, published studies may result distorted meta-analysis because of studies with statistically significant result are more likely to be published than the insignificant one which might induce overestimation of the treatment efficacy in the review (Dickersin et al., 1987). Apart from that, lack of sensitivity analyses and publication bias assessment is one of the major limitations of the study. A limited number of RCTs (n=3) comparing the effectiveness of Metformin and Thiazolidinedione. Women with Polycystic ovary syndrome in all included RCTs were recruited from three different hospitals in Europe which may constrain the potential applicability of the review results if any ethnic variation exists.

## 6. Conclusion

In conclusion, this systematic review represents an equal effect of Metformin and Thiazolidinediones on endothelial function among women with PCOS, though most of the individual studies shows beneficial outcome. Even, the Placebo-controlled clinical trials demonstrated positive effects of both of Metformin and Thiazolidinedione on vascular function. Since, both insulin sensitizers revealed numerous effects at molecular level along with improved insulin sensitivity, therefore this review intended to show the comparative effectiveness of Metformin and Thiazolidinediones on endothelial function in Polycystic ovary syndrome. Although, this review resulting comparable effects of Metformin and Thiazolidinediones on vascular endothelial function, it is noteworthy that all the trials conducted on a very narrow scale which might limit the statistical accuracy.

## Data Availability

Since this is a systematic review, hence data was collected from the primary studies included in this review as per predetermined eligibility criteria.

## Acknowledgements

The authors wish to thank Dr. Mzwandile A Mabhala and Dr. Alan Massey of University of Chester for their support in preparing manuscript.

